# Comparison of Mental Health Symptoms Prior to and During COVID-19 among Patients with Systemic Sclerosis from Four Countries: A Scleroderma Patient-centered Intervention Network (SPIN) Cohort Study

**DOI:** 10.1101/2020.06.13.20128694

**Authors:** Brett D. Thombs, Linda Kwakkenbos, Richard S. Henry, Marie-Eve Carrier, Scott Patten, Sami Harb, Angelica Bourgeault, Lydia Tao, Susan J. Bartlett, Luc Mouthon, John Varga, Andrea Benedetti, for the SPIN Patient Advisors, SPIN Investigators

**Affiliations:** Lady Davis Institute for Medical Research, Jewish General Hospital, 4333 Chemin de la Côte-Sainte-Catherine, Montreal, Quebec, H3T 1E4, Canada; Department of Psychiatry, McGill University, 1033 Pine Avenue West, Montreal, Quebec, H3A 1A1, Canada; Department of Epidemiology, Biostatistics, and Occupational Health, McGill University, 1020 Pine Avenue West, Montreal, Quebec, H3A 1A2, Canada; Department of Medicine, McGill University, 1001 Decarie Boulevard, Montreal, Quebec, H4A 3J1, Canada; Department of Psychology, McGill University, 2001 McGill College Avenue, Montreal, Quebec, H3A 1G1, Canada; Department of Educational and Counselling Psychology, McGill University, 3700 McTavish Street, Montreal, Quebec, H3A 1Y2, Canada; Biomedical Ethics Unit, McGill University, 3647 Peel Street, Montreal, Quebec, H3A 1X1, Canada; Department of Clinical Psychology, Behavioural Science Institute, Radboud University, Montessorilaan 3, 6525 HR, Nijmegen, the Netherlands; Department of Community Health Sciences, University of Calgary, 3280 Hospital Drive NW, Calgary, Alberta, T2N 4Z6, Canada; Hotchkiss Brain Institute, University of Calgary, 3330 Hospital Drive NW, Calgary, Alberta, T2N 4N1, Canada; O’Brien Institute for Public Health, University of Calgary, 3280 Hospital Drive NW, Calgary, Alberta, T2N 4Z6, Canada; Research Institute of the McGill University Health Centre, Centre for Outcomes Research and Evaluation, 5252 de Maisonneuve #3D.57, Montreal, Quebec, H4A 2S5, Canada; Université Paris Descartes, Assistance Publique-Hôpitaux de Paris, 3 Avenue Victoria, 75004 Paris, France; Service de Médecine Interne, Centre de Reference Maladies Systémiques Autoimmunes Rares d’Ile de France, Hôpital Cochin, Assistance Publique - Hôpitaux de Paris, 27 Rue du Faubourg Saint-Jacques, 75014 Paris, France; Northwestern Scleroderma Program, Feinberg School of Medicine, Northwestern University, 675 North St. Clair, Chicago, Illinois, 60611, USA; Respiratory Epidemiology and Clinical Research Unit, McGill University Health Centre, 5252 boulevard de Maisonneuve, Montreal, Quebec, H4A 3S5, Canada; SPIN Patient Advisors include Catherine Fortuné, Scleroderma Society of Ontario, Hamilton, Ontario, Canada; Amy Gietzen, Scleroderma Foundation, Tri-State Chapter, Binghamton, New York, USA; Geneviève Guillot, Sclérodermie Québec, Longueuil, Quebec, Canada; Nancy Lewis, Toronto, Ontario, Canada; Michelle Richard, Scleroderma Atlantic, Halifax, Nova Scotia, Canada; Maureen Sauvé, Scleroderma Society of Ontario, Hamilton, Ontario, Canada; Joep Welling, NVLE Dutch patient organization for systemic autoimmune diseases, Utrecht, The Netherlands; Kim Fligelstone, Scleroderma & Raynaud’s UK, London, United Kingdom; Karen Gottesman, Scleroderma Foundation, Los Angeles, California, USA; Catarina Leite, University of Minho, Braga, Portugal; Elisabet Pérez, Asociación Española de Esclerodermia, Madrid, Spain.; SPIN Investigators include Murray Baron, McGill University, Montreal, Quebec, Canada; Vanessa Malcarne, San Diego State University, San Diego, California, USA; Maureen D. Mayes, University of Texas McGovern School of Medicine, Houston, Texas, USA; Warren R. Nielson, St. Joseph’s Health Care, London, Ontario, Canada; Robert Riggs, Scleroderma Foundation, Danvers, Massachusetts, USA; Shervin Assassi, University of Texas McGovern School of Medicine, Houston, Texas, USA; Carolyn Ells, McGill University, Montreal, Quebec, Canada; Cornelia van den Ende, Sint Maartenskliniek, Nijmegen, The Netherlands; Tracy Frech, University of Utah, Salt Lake City, Utah, USA; Daphna Harel, New York University, New York, New York, USA; Monique Hinchcliff, Yale School of Medicine, New Haven, Connecticut, USA; Marie Hudson, McGill University, Montreal, Quebec, Canada; Sindhu R. Johnson, Toronto Scleroderma Program, Mount Sinai Hospital, Toronto Western Hospital, and University of Toronto, Toronto, Ontario, Canada; Maggie Larche, McMaster University, Hamilton, Ontario, Canada; Christelle Nguyen, Université Paris Descartes, and Assistance Publique - Hôpitaux de Paris, Paris, France; Janet Pope, University of Western Ontario, London, Ontario, Canada; François Rannou, Université Paris Descartes, and Assistance Publique - Hôpitaux de Paris, Paris, France; Tatiana Sofia Rodriguez Reyna, Instituto Nacional de Ciencias Médicas y Nutrición Salvador Zubiràn, Mexico City, Mexico; Anne A. Schouffoer, Leiden University Medical Center, Leiden, The Netherlands; Maria E. Suarez-Almazor, University of Texas MD Anderson Cancer Center, Houston, Texas, USA; Christian Agard, Centre Hospitalier Universitaire - Hôtel-Dieu de Nantes, Nantes, France; Alexandra Albert, Université Laval, Quebec, Quebec, Canada; Elana J. Bernstein, Columbia University, New York, New York, USA; Sabine Berthier, Centre Hospitalier Universitaire Dijon Bourgogne, Dijon, France; Benjamin Chaigne, Assistance Publique - Hôpitaux de Paris, Hôpital Cochin, Paris, France; Lorinda Chung, Stanford University, Stanford, California, USA; Chase Correia, Northwestern University, Chicago, Illinois, USA; Christopher Denton, Royal Free London Hospital, London, UK; Robyn Domsic, University of Pittsburgh, Pittsburgh, Pennsylvania, USA; James V. Dunne, St. Paul’s Hospital and University of British Columbia, Vancouver, British Columbia, Canada; Bertrand Dunogue, Assistance Publique - Hôpitaux de Paris, Hôpital Cochin, Paris, France; Dominique Farge-Bancel, Assistance Publique - Hôpitaux de Paris, Hôpital St-Louis, Paris, France; Paul R. Fortin, CHU de Québec - Université Laval, Quebec, Quebec, Canada; Jessica Gordon, Hospital for Special Surgery, New York City, New York, USA; Brigitte Granel-Rey, Aix Marseille Université, and Assistance Publique - Hôpitaux de Marseille, Hôpital Nord, Marseille, France; Pierre-Yves Hatron, Centre Hospitalier Régional Universitaire de Lille, Hôpital Claude Huriez, Lille, France; Ariane L Herrick, University of Manchester, Salford Royal NHS Foundation Trust, Manchester, UK; Sabrina Hoa, Centre hospitalier de l’université de Montréal – CHUM, Montreal, Quebec, Canada; Niall Jones, University of Alberta, Edmonton, Alberta, Canada; Artur Jose de B. Fernandes, Université de Sherbrooke, Sherbrooke, Quebec, Canada; Suzanne Kafaja, University of California, Los Angeles, California, USA; Nader Khalidi, McMaster University, Hamilton, Ontario, Canada; David Launay, Centre Hospitalier Régional Universitaire de Lille, Hôpital Claude Huriez, Lille, France; Joanne Manning, Salford Royal NHS Foundation Trust, Salford, UK; Isabelle Marie, CHU Rouen, Hôpital de Bois-Guillaume, Rouen, France; Maria Martin, Servicio de Reumatologia del Hospital 12 de Octubre, Madrid, SpainFrançois Maurier, Hôpitaux Privés de Metz, Hôpital Belle-Isle, Metz, France; Arsene Mekinian, Assistance Publique - Hôpitaux de Paris, Hôpital St-Antoine, Paris, France; Sheila Melchor, Servicio de Reumatologia del Hospital 12 de Octubre, Madrid, Spain; Mandana Nikpour, St Vincent’s Hospital and University of Melbourne, Melbourne, Victoria, Australia; Louis Olagne, Centre Hospitalier Universitaire Gabriel-Montpied, Clermont-Ferrand, France; Susanna Proudman, Royal Adelaide Hospital and University of Adelaide, Adelaide, South Australia, Australia; Alexis Régent, Assistance Publique - Hôpitaux de Paris, Hôpital Cochin, Paris, France; Sébastien Rivière, Assistance Publique - Hôpitaux de Paris, Hôpital St-Antoine, Paris, France; David Robinson, University of Manitoba, Winnipeg, Manitoba, Canada; Esther Rodriguez, Servicio de Reumatologia del Hospital 12 de Octubre, Madrid, Spain; Sophie Roux, Université de Sherbrooke, Sherbrooke, Quebec, Canada; Vincent Sobanski, Centre Hospitalier Régional Universitaire de Lille, Hôpital Claude Huriez, Lille, France; Virginia Steen, Georgetown University, Washington, DC, USA; Evelyn Sutton, Dalhousie University, Halifax, Nova Scotia, Canada; Carter Thorne, Southlake Regional Health Centre, Newmarket, Ontario, Canada; Pearce Wilcox, St. Paul’s Hospital and University of British Columbia, Vancouver, British Columbia, Canada; Canada; Mara Cañedo Ayala, Jewish General Hospital, Montreal, Quebec, Canada; Andrea Carboni-Jiménez, Jewish General Hospital, Montreal, Quebec, Canada; Maria Gagarine, Jewish General Hospital, Montreal, Quebec, Canada; Julia Nordlund, Jewish General Hospital, Montreal, Quebec, Canada; Nora Østbø, Jewish General Hospital, Montreal, Quebec, Canada; Danielle B. Rice, Jewish General Hospital, Montreal, Quebec, Canada; Kimberly A. Turner, Jewish General Hospital, Montreal, Quebec, Canada. Nicole Culos-Reed, University of Calgary, Alberta, Canada; Laura Dyas, Scleroderma Foundation Michigan Chapter, Southfield, Michigan, USA; Ghassan El-Baalbaki, Université du Québec à Montréal, Montreal, Quebec, Canada; Shannon Hebblethwaite, Concordia University, Montreal, Quebec, Canada; Laura Bustamante, Concordia University, Montreal, Quebec, Canada; Delaney Duchek, University of Calgary, Alberta, Canada; Kelsey Ellis, University of Calgary, Alberta, Canada.

## Abstract

**Background:** No studies have reported comparisons of mental health symptoms prior to and during COVID-19 in vulnerable populations. Objectives were to compare anxiety and depression symptoms among people with a pre-existing medical condition, the autoimmune disease systemic sclerosis (SSc; scleroderma), including continuous change scores, proportion with change ≥ 1 minimal clinically important difference (MCID), and factors associated with changes, including country.

**Methods:** Pre-COVID-19 Scleroderma Patient-centered Intervention Network Cohort data were linked to COVID-19 data collected April 9 to April 27, 2020. Anxiety symptoms were assessed with the PROMIS Anxiety 4a v1.0 scale (MCID = 4 points) and depression symptoms with the Patient Health Questionnaire-8 (MCID = 3 points). Multiple linear and logistic regression were used to assess factors associated with continuous change and change **≥** 1 MCID.

**Findings:** Among 435 participants (Canada = 98; France = 159; United Kingdom = 50; United States = 128), mean anxiety symptoms increased 4.9 points (95% confidence interval [CI] 4.0 to 5.7). Depression symptom change was negligible (0.3 points; 95% CI −0.7 to 0.2). Compared to France, adjusted scores from the United States and United Kingdom were 3.8 points (95% CI 1.7 to 5.9) and 2.9 points higher (95% CI 0.0 to 5.7); scores for Canada were not significantly different. Odds of increasing by ≥ 1 MCID were twice as high for the United Kingdom (2.0, 95% CI 1.0 to 4.2) and United States (1.9, 95% CI 1.1 to 3.2). Participants who used mental health services pre-COVID had adjusted increases 3.7 points (95% CI 1.7 to 5.7) less than other participants.

**Interpretation:** Anxiety symptoms, but not depression symptoms, increased dramatically during COVID-19 among people with a pre-existing medical condition. Increase was larger in the United Kingdom and United States than in Canada and France but substantially less for people with pre-COVID-19 mental health treatment.

**RESEARCH IN CONTEXT:** *Evidence before this study:* We referred to a living systematic review that is evaluating mental health changes from pre-COVID-19 to COVID-19 by searching 7 databases, including 2 Chinese language databases, plus preprint servers, with daily updates (https://www.depressd.ca/covid-19-mental-health). As of June 13, 2020, only 5 studies had compared mental health symptoms prior to and during COVID-19. In 4 studies of university students, there were small increases in depression or general mental health symptoms but minimal or no increases in anxiety symptoms. A general population study from the United Kingdom reported a small increase in general mental health symptoms but did not differentiate between types of symptoms. No studies have reported changes from pre-COVID-19 among people vulnerable due to pre-existing medical conditions. No studies have compared mental health changes between countries, despite major differences in pandemic responses.

*Added value of this study:* We evaluated changes in anxiety and depression symptoms among 435 participants with the autoimmune condition systemic sclerosis and compared results from Canada, France, the United Kingdom, and the United States. To our knowledge, this is the first study to compare mental health symptoms prior to and during COVID-19 in any vulnerable population. These are the first data to document the substantial degree to which anxiety symptoms have increased and the minimal changes in depression symptoms among vulnerable individuals. It is also the first study to examine the association of symptom changes with country of residence and to identify that people receiving pre-COVID-19 mental health services may be more resilient and experience less substantial symptom increases than others.

*Implications of all the available evidence:* Although this was an observational study, it provided evidence that vulnerable people with a pre-existing medical condition have experienced substantially increased anxiety symptoms and that these increases appear to be associated with where people live and, possibly, different experiences of the COVID-19 pandemic across countries. By comparing with evidence from university samples, which found that depression symptoms were more prominent, these data underline the need for accessible interventions tailored to specific needs of different populations. They also suggest that mental health treatments may help people to develop skills or create resilience, which may reduce vulnerability to major stressors such as COVID-19.

## INTRODUCTION

The SARS-CoV-2 coronavirus disease (COVID-19) pandemic has caused more than 400,000 deaths and has had devastating health, social, political, and economic consequences worldwide. There are expected to be serious mental health implications during and beyond the initial outbreak, but their degree and nature are not well understood.^1-2^

Many cross-sectional studies report percentages of participants above cutoff thresholds on mental health symptom questionnaires during COVID-19. Such percentages, however, vary substantially across otherwise similar populations even in normal times.^3^ Furthermore, they tend to dramatically overestimate prevalence obtained from validated methods, and there is too much heterogeneity to correct for differences statistically.^4,5^ Thus, studies that directly evaluate changes are needed.

Based on a living systematic review,^3,6^ as of June 13, 2020, only 5 studies had compared mental health prior to and during COVID-19. Four studies of university students suggest small increases in depression but minimal or no increases in anxiety. A United Kingdom general population study found small increases in general mental health symptoms but did not differentiate between anxiety and depression symptoms. No studies have evaluated mental health changes among people at risk of COVID-19 complications due to pre-existing medical conditions. Furthermore, despite important differences in pandemic responses across countries, no studies have compared mental health changes between countries.

People with the autoimmune disease systemic sclerosis (SSc; scleroderma) are representative of patients with pre-existing medical conditions that put them at risk during COVID-19. More than 40% have interstitial lung disease, many are frail, and use of immunosuppressant drugs is common.^7,8^ The Scleroderma Patient-centered Intervention Network (SPIN) Cohort routinely collects mental health outcomes at 3-to 6-month intervals.^8-10^ The SPIN COVID-19 Cohort was initiated to collect data during COVID-19 and allows comparison of mental health symptoms prior to and during COVID-19 for participants enrolled in both cohorts.

Our objective was to compare anxiety (PROMIS Anxiety 4a v1.0 scale^11,12^) and depression (Patient Health Questionnaire-8 [PHQ-8]^13^) symptoms before and after onset of COVID-19 among people with SSc, including (1) continuous score changes; (2) proportion with change scores of at least one minimal clinically important difference (MCID); (3) proportion initially under a cutoff threshold who changed by at least 1 MCID and reached the threshold; and (4) factors associated with changes, including country, comparing results from Canada, France, the United Kingdom, and the United States.

## METHODS

This was a longitudinal study that linked pre-COVID-19 data from the SPIN Cohort^8-10^ to data collected during the baseline assessment of the separate SPIN COVID-19 Cohort between April 9, 2020 and April 27, 2020 using the same measurement scales. Person-level, deterministic linking was used with participant email addresses as the indentifier. The full protocol for the SPIN COVID-19 Cohort and the present study is available online (https://osf.io/kbncx/).

### Participants and Procedure

SPIN Cohort participants must be aged ≥ 18 years and meet 2013 American College of Rheumatology/European League Against Rheumatism criteria for SSc, verified by a SPIN physician.^14^ The SPIN Cohort is a convenience sample.^8^ Eligible participants are recruited at 47 SPIN sites^10^ in Canada, the United States, the United Kingdom, France, Spain, Mexico, and Australia during regular medical visits. Site personnel submit an online medical form to enrol participants, after which participants receive an email with instructions to activate their SPIN account and complete measures via the Cohort online portal in English, French, or Spanish. Assessments are completed at 3-month intervals. SPIN Cohort participants provide informed consent for cohort participation and for contact about additional SPIN studies.

From April 9 to April 27, 2020, SPIN Cohort participants who complete measures in English or French were invited by email and popups during SPIN Cohort online assessments to enrol in the SPIN COVID-19 Cohort. SPIN Cohort participants included in the present study (1) were from Canada, the United States, the United Kingdom, and France; (2) completed the PROMIS Anxiety 4a v1.0 scale^11,12^ in English or French between July 1, 2019 and December 31, 2019, when China reported cases of pneumonia later identified as related to COVID-19 to the World Health Organization^15^; and (3) enrolled in the SPIN COVID-19 Cohort and completed baseline measures. SPIN COVID-19 measures were collected using the *Qualtrics* online survey package.

The SPIN (#MP-05-2013-150) and SPIN COVID-19 (#2021-2286) Cohorts were approved by the Research Ethics Committee of the Centre intégré universitaire de santé et de services sociaux du Centre-Ouest-de-l’Île-de-Montréal. The SPIN Cohort was also approved by ethics committees of SPIN sites.

### Measures

Physician-reported SPIN Cohort data included sex, age, body mass index, time since SSc diagnosis, SSc disease subtype (limited, diffuse, sine scleroderma), presence of interstitial lung disease, and presence of overlap syndromes (systemic lupus erythematosus, rheumatoid arthritis, Sjögrens syndrome, idiopathic inflammatory myopathy, primary biliary cirrhosis, autoimmune thyroid disease). Pre-COVID-19 patient-reported data included race or ethnicity, employment status, health professional visit about mental health in previous 3 months, interference of breathing problems in daily activities (single item, past-week, 0-10 severity), the PROMIS Physical Function 4a v1.0 scale,^11,12^ the PROMIS Anxiety 4a v1.0 scale,^11,12^ and the PHQ-8.^13^ Patient-reported data during COVID-19 included immunosuppressant drug use, COVID-19 positive test status, financial resource adequacy (Consumer Financial Protection Bureau Financial Well-Being Scale^16^), and anxiety and depression symptoms. Details are available in the study protocol.

#### Anxiety Symptoms

The PROMIS Anxiety 4a v1.0 scale^11,12^ includes 4 items asking participants, in the past 7 days, how often: (1) “I felt fearful”; (2) “I found it hard to focus on anything other than my anxiety”; (3) “My worries overwhelmed me”; and (4) “I felt uneasy”. Items are scored 1-5 with response options “never” to “always”. Higher scores represent more anxiety. Raw scores are converted into T-scores standardized from the general US population (mean = 50, standard deviation = 10). A change of 4.0 T-score points was selected to represent the MCID^17^ and a threshold for identifying people with at least moderate symptoms of T-score ≥ 60.^11^ PROMIS Anxiety 4a v1.0 has been validated in SSc^18,19^ and is included in all 3-month SPIN Cohort assessments.

#### Depressive Symptoms

The eight-item PHQ-8^13^ measures depressive symptoms over the last 2 weeks with item scores from 0 (not at all) to 3 (nearly every day) and higher scores representing more depression. The MCID has been estimated to be 3.0 points,^20^ and a threshold of ≥ 10 is commonly used to identify people who may have depression.^21^ The PHQ-8, which is assessed every 6 months in the SPIN Cohort, performs equivalently to the PHQ-9,^22^ which has been shown to be valid in SSc.^23^

### Statistical Analyses

Descriptive statistics are presented as mean (standard deviation) for continuous variables and numbers (percentages) for categorical variables. Changes in anxiety and depression symptoms were described: (1) continuously with T-scores or raw scores, in terms of MCIDs, and with a Hedges g standardized mean difference effect size, all with 95% confidence intervals (CIs); (2) as the proportion of participants whose symptoms worsened or improved, separately, by at least 1 MCID, with 95% CIs; and (3) as the proportion initially below a T-score of 60 on the PROMIS Anxiety 4a v1.0^12^ or a score of 10 on the PHQ-8^22^ who increased by at least 1 MCID and reached the threshold score, with 95% CIs. For proportions, 95% CIs were generated based on Agresti and Coull’s approximate method for binomial proportions.^24^

We conducted two sets of sensitivity analyses for symptom changes. First, for both anxiety and depressive symptoms, we compared change to scores from assessments done between January 1 and June 30, 2019 in order to determine if seasonal variations may have influenced our main findings. Second, since the PROMIS Anxiety 4a v1.0 is administered in the SPIN Cohort every 3 months and the PHQ-8 every 6 months, to evaluate whether differences in change between anxiety and depression symptoms may have been due to different assessment points between July 1 and December 31, 2019, we evaluated changes only including assessments done at the same timepoint.

We evaluated the association of sociodemographic characteristics, medical characteristics, and COVID-19 variables with continuous changes in anxiety and depression symptoms via multivariable linear regression and with a change of ≥1 MCID with multivariable logistic regression. All variables were selected *a priori* and entered simultaneously. For continuous variables, we assessed linearity via restricted cubic splines. Missing data were dealt with using multiple imputation via chained equations with 20 imputations. Variables entered in models included male sex (reference female), age (continuous), non-White race or ethnicity (reference White), education years (continuous), living alone (reference living with others), country (reference France), working part- or full-time (reference not working), time since SSc diagnosis (continuous), diffuse subtype (reference limited or sine scleroderma), interstitial lung disease presence, interference from breathing problems (continuous), overweight or obese (reference normal body mass index or less), overlap syndrome presence, PROMIS Physical Function 4a v1.0 (continuous), immunosuppressant drug use presence, use of mental health services pre-COVID, and financial resource adequacy (continuous).

All analyses were conducted using Stata (Version 13) with 2-sided statistical tests and p < 0.05 significance level.

### Changes to Protocol

Changes included exclusion of participants from Australia, because only 10 would have been eligible; removal of COVID-19 infection from model covariates, since only 4 participants reported a positive test; and addition of sensitivity analyses.

### Role of the Funding Source

Funders had no role in any aspect of study design; data collection, analysis and interpretation; manuscript drafting; or the decision to submit for publication. The corresponding author had access to all data and final responsibility for the decision to submit for publication.

## RESULTS

### Participants

There were 435 SPIN Cohort participants from Canada (N = 98), France (N = 159), the United Kingdom (N = 50), and the United States (N = 128) who enrolled in the SPIN COVID-19 Cohort and were included in the present study. See Figure 1 for participant flow and Supplementary Table 1 for number of participants from recruitment sites. Table 1 shows participant characteristics. Mean age was 56.9 years, and 88.5% of participants were female. Mean time since SSc diagnosis was 12.1 years; 39.8% had diffuse disease subtype, 35.2% had interstitial lung disease, and 48.1% were using immunosuppressant drugs. Participant characteristics were similar for most variables across countries.

**Table 1.** Participant Characteristics for the Full Sample and by Country.

|  | Overall<br>(N=435) | Canada<br>(N=98) | France<br>(N=159) | United Kingdom<br>(N=50) | United States<br>(N=128) |
| --- | --- | --- | --- | --- | --- |
| Variable | Mean (SD) or<br>N (%) | Mean (SD) or<br>N (%) | Mean (SD) or<br>N (%) | Mean (SD) or<br>N (%) | Mean (SD) or<br>N (%) |
| **Sociodemographics** |  |  |  |  |  |
| Age in years, *mean (SD)* | 56.9 (12.6) | 57.4 (11.5) | 53.8 (14.8) | 59.2 (10.5) | 59.5 (10.4) |
| Male sex, *N (%)* | 50 (11.5) | 11 (11.2) | 16 (10.1) | 4 (8.0) | 19 (14.8) |
| Education in years, *mean (SD)* | 15.6 (3.7)<sup>a</sup> | 15.6 (3.2)<sup>b</sup> | 15.1 (4.6)<sup>c</sup> | 14.8 (3.3)<sup>d</sup> | 16.4 (2.8) |
| Living alone, *N (%)* | 80 (18.7)<sup>e</sup> | 14 (14.4)<sup>f</sup> | 33 (21.6)<sup>g</sup> | 11 (22.0) | 22 (17.3)<sup>h</sup> |
| Race/ethnicity, *N (%)* |  |  |  |  |  |
| White | 360 (83.0)<sup>i</sup> | 87 (88.8) | 122 (77.2)<sup>c</sup> | 45 (90.0) | 106 (82.8) |
| Black | 38 (8.8)<sup>i</sup> | 1 (1.0) | 24 (15.2)<sup>c</sup> | 4 (8.0) | 9 (7.0) |
| Other | 36 (8.3)<sup>i</sup> | 10 (10.2) | 12 (7.6)<sup>c</sup> | 1 (2.0) | 13 (10.2) |
| Working part- or full-time, *N (%)* | 197 (45.4)<sup>i</sup> | 44 (44.9) | 74 (46.8)<sup>c</sup> | 21 (42.0) | 58 (45.3) |
| **Medical characteristics** |  |  |  |  |  |
| Body Mass Index, *N (%)* |  |  |  |  |  |
| Underweight or normal (< 25)<sup>j</sup> | 254 (58.4) | 53 (54.1) | 107 (67.3) | 26 (52.0) | 68 (53.1) |
| Overweight (25 to <30) | 110 (25.3) | 19 (19.4) | 36 (22.6) | 16 (32.0) | 39 (30.5) |
| Obese (≥ 30.0) | 71 (16.3) | 26 (26.5) | 16 (10.1) | 8 (16.0) | 21 (16.4) |
| Time since diagnosis in years, *mean (SD)* | 12.1 (7.8)<sup>k</sup> | 12.4 (9.7)<sup>b</sup> | 11.1 (7.2)<sup>c</sup> | 13.3 (7.8)<sup>l</sup> | 12.7 (6.6)<sup>m</sup> |
| Diffuse disease subtype, *N (%)* | 173 (39.8) | 40 (40.8) | 56 (35.2) | 17 (34.0) | 60 (46.9) |
| Presence of interstitial lung disease, *N* (%) | 148 (35.2)<sup>n</sup> | 28 (30.4)<sup>o</sup> | 57 (36.3)<sup>p</sup> | 14 (28.6)<sup>d</sup> | 49 (39.8)<sup>q</sup> |
| Presence of overlap syndrome, *N* (%) | 100 (24.0)<sup>r</sup> | 22 (23.9)<sup>o</sup> | 28 (17.8)<sup>p</sup> | 23 (46.0) | 27 (22.9)<sup>s</sup> |
| Immunosuppressant drug use during COVID-19, *N* (%) | 209 (48.1) | 44 (44.9) | 66 (41.5) | 31 (62.0) | 68 (53.1) |
| Pre-COVID-19 use of mental health services, *N* (%) | 92 (21.2%) | 24 (24.5) | 31 (19.5) | 9 (18.0) | 28 (21.9) |
| COVID-19 infection confirmed by test, *N* (%) | 4 (0.9) | 0 (0.0) | 4 (2.5) | 0 (0.0) | 0 (0.0) |
| **Patient-reported outcomes** |  |  |  |  |  |
| Patient-reported interference from breathing problems pre-COVID, *mean* (*SD*) | 2.5 (2.9)<sup>t</sup> | 2.9 (3.0)<sup>f</sup> | 2.6 (3.0) | 2.7 (3.2)<sup>d</sup> | 2.1 (2.5)<sup>h</sup> |
| PROMIS Physical Function 4a v1.0 pre-COVID, *mean* (*SD*) | 43.6 (8.7) | 42.4 (8.5) | 44.6 (9.2) | 43.1 (8.9) | 43.4 (8.0) |
| Adequacy of financial resources score, *mean* (*SD*) | 13.0 (4.8)<sup>u</sup> | 13.1 (5.3)<sup>f</sup> | 12.7 (5.0)<sup>v</sup> | 13.2 (4.1) | 13.2 (4.3) |
| **Number of days since completion of pre-COVID measures** |  |  |  |  |  |
| PROMIS Anxiety 4a v1.0, *mean* (*SD*) | 163.1 (37.4) | 154.1 (29.3) | 163.4 (38.5) | 167.2 (37.4) | 168.2 (40.6) |
| Patient Health Questionnaire-8, *mean* (*SD*) | 197.0 (54.0)<sup>w</sup> | 189.2 (57.7)<sup>x</sup> | 197.4 (52.5)<sup>y</sup> | 197.3 (50.8)<sup>z</sup> | 203.0 (54.0)<sup>aa</sup> |

<sup>a</sup>N=431, <sup>b</sup>N=96, <sup>c</sup>N=158, <sup>d</sup>N=49, <sup>e</sup>N=427, <sup>f</sup>N=97, <sup>g</sup>N=153, <sup>h</sup>N=127, <sup>i</sup>N=434, <sup>j</sup>Because N underweight = 24, underweight and normal were combined, <sup>k</sup>N=422, <sup>l</sup>N=44, <sup>m</sup>N=124, <sup>n</sup>N=421, <sup>o</sup>N=92, <sup>p</sup>N=157, <sup>q</sup>N=123, <sup>r</sup>N=417, <sup>s</sup>N=118, <sup>t</sup>N=432, <sup>u</sup>N=430, <sup>v</sup>N=155, <sup>w</sup>N=388, <sup>x</sup>N=91, <sup>y</sup>N=146, <sup>z</sup>N=43, <sup>aa</sup>N=108.

**Figure 1.**
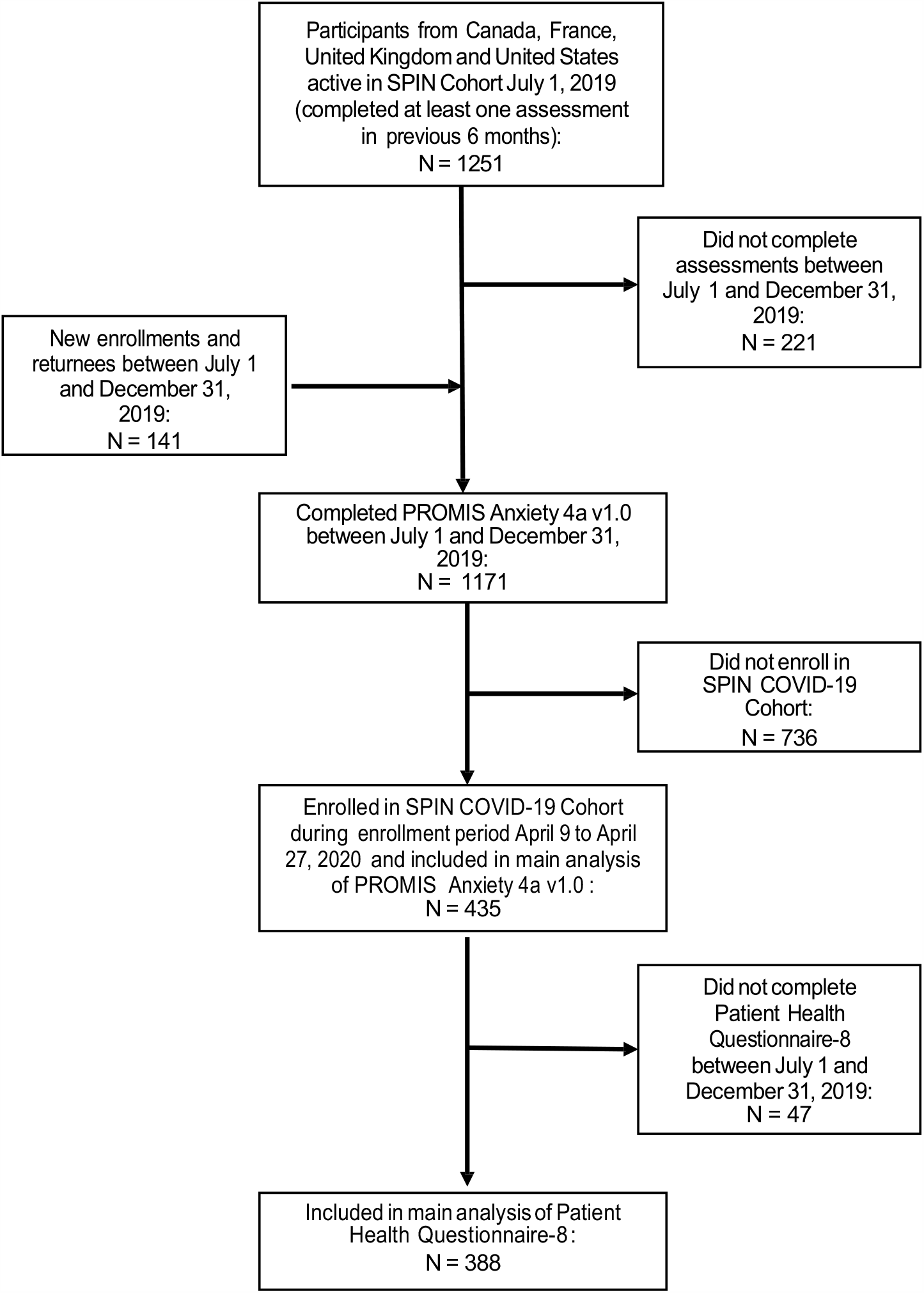
Flow diagram of participant enrollment and inclusion.

### Comparison of Symptoms of Anxiety and Depression Prior to and During COVID-19

As shown in Table 2, anxiety symptoms increased more than a full MCID (4.9 points, 95% CI 4.0 to 5.7). Increases by country were 3.1 points (95% CI 1.7 to 4.6) for France, 4.4 points for Canada (95% CI 2.7 to 6.0), 6.2 points for the United Kingdom (95% CI 4.0 to 8.3), and 6.9 points for the United States (95% CI 5.4 to 8.5). The percentage of participants with ≥ 1 MCID increase was 42.8% (95% CI 35.3% to 50.5%) for France, 46.9% for Canada (95% CI 37.4% to 56.7%), 59.4% (95% CI 50.7% to 67.5%) for the United States, and 64.0% (95% CI 50.1% to 75.9%) for the United Kingdom. A similar increase in anxiety was seen compared to pre-COVID-19 anxiety symptoms assessed January 1, 2019 to June 30, 2019 (N = 392; see Supplementary Table 2).

**Table 2.** **Change in Symptoms of PROMIS Anxiety 4a v1**.**0 Pre-COVID-19 to COVID-19 (Higher Scores = Greater Anxiety)**

|  | Full sample | Canada | France | United Kingdom | United States |
| --- | --- | --- | --- | --- | --- |
|  | N = 435 | N = 98 | N = 159 | N = 50 | N = 128 |
| **Change in Score:** |  |  |  |  |  |
| Pre-COVID-19, *mean* | 52.7 (10.4) | 54.3 (10.7) | 53.6 (10.8) | 53.4 (9.9) | 50.0 (9.5) |
| (*SD*) |  |  |  |  |  |
| Post-COVID-19, *mean* | 57.5 (8.8) | 58.6 (8.2) | 56.7 (9.6) | 59.6 (8.1) | 56.9 (8.3) |
| (*SD*) |  |  |  |  |  |
| Mean change (95% CI) | 4.9 (4.0 to 5.7) | 4.4 (2.7 to 6.0) | 3.1 (1.7 to 4.6) | 6.2 (4.0 to 8.3) | 6.9 (5.4 to 8.5) |
| Hedges' g (95% CI) | 0.51 (0.37 to 0.64) | 0.46 (0.17 to 0.74) | 0.30 (0.08 to 0.52) | 0.67 (0.27 to 1.08) | 0.77 (0.51 to 1.02) |
| **Change of ≥ 1 MCID:** |  |  |  |  |  |
| N worsening (% , 95% CI) | 222 | 46 | 68 | 32 | 76 |
|  | (51.0%, 46.4% to 55.7%) | (46.9%, 37.4% to 56.7%) | (42.8%, 35.3% to 50.5%) | (64.0%, 50.1% to 75.9%) | (59.4%, 50.7% to 67.5%) |
| N improving (% , 95% CI) | 55 | 14 | 29 | 5 | 7 |
|  | (12.6%, 9.8% to 16.1%) | (14.3%, 8.7% to 22.6%) | (18.2%, 13.0% to 25.0%) | (10.0%, 4.4% to 21.4%) | (5.5%, 2.7% to 10.9%) |
| **Change of ≥ 1 MCID and reaches cutoff threshold<sup>a</sup>** |  |  |  |  |  |
| N of N initially < cutoff | 81 of 324 | 15 of 72 | 22 of 108 | 15 of 36 | 29 of 108 |
| worsening (% , 95% CI) | (25.0%, 20.6% to 30.0%) | (20.8%, 13.1% to 31.6%) | (20.4%, 13.9% to 28.9%) | (41.7%, 27.1% to 57.8%) | (26.9%, 19.4% to 35.9%) |

MCID = minimal clinically important difference. <sup>a</sup> Cutoff threshold = ≥ 60 for PROMIS Anxiety 4a v1.0

As shown in Table 3, among 388 participants who completed the PHQ-8 in the last 6 months of 2019, changes in depressive symptoms were minimal (reduction of 0.3 points, 95% CI −0.7 to 0.2). As shown in Supplementary Table 2, this result was unchanged when including only assessments done on the same day as the included PROMIS Anxiety 4a v1.0 assessments (N = 223) and compared to results from assessments done in the first 6 months of 2019 (N = 352).

**Table 3.** Change in Symptoms of Depression Pre-COVID-19 to COVID-19.

|  | Full sample | Canada | France | United Kingdom | United States |
| --- | --- | --- | --- | --- | --- |
|  | N = 388 | N = 91 | N = 146 | N = 43 | N = 108 |
| **Change in Score:** |  |  |  |  |  |
| Pre-COVID-19, *mean* | 6.7 (5.7) | 7.2 (6.1) | 7.4 (5.9) | 7.2 (5.9) | 5.3 (4.9) |
| (*SD*) |  |  |  |  |  |
| Post-COVID-19, *mean* | 6.4 (5.4) | 7.2 (5.9) | 6.2 (5.2) | 7.5 (5.8) | 5.7 (5.2) |
| (*SD*) |  |  |  |  |  |
| Mean change (95% CI) | -0.3 (-0.7 to 0.2) | 0.0 (-1.04 to 1.0) | -1.1 (-1.9 to -0.4) | 0.3 (-1.1 to 1.7) | 0.4 (-0.3 to 1.1) |
| Hedges' g (95% CI) | -0.05 (-0.19 to 0.09) | 0.00 (-0.29 to 0.29) | -0.20 (-0.43 to 0.03) | 0.05 (-0.38 to 0.48) | 0.08 (-0.19 to 0.35) |
| **Change of ≥ 1 MCID:** |  |  |  |  |  |
| N worsening (% , 95% CI) | 82 | 23 | 24 | 11 | 24 |
|  | (21.1%, 17.4% to 25.5%) | (25.3%, 17.5% to 35.1%) | (16.4%, 11.3% to 23.3%) | (25.6, 14.9% to 40.2%) | (22.2%, 15.4% to 30.9%) |
| N improving (% , 95% CI) | 80 | 18 | 40 | 9 | 13 |
|  | (20.6%, 16.9% to 24.9%) | (19.8%, 12.9% to 29.1%) | (27.4%, 20.8% to 35.1%) | (20.9%, 11.4% to 35.2%) | (12.0%, 7.2% to 19.5%) |
| **Change of ≥ 1 MCID and reaches cutoff threshold<sup>a</sup>** |  |  |  |  |  |
| N of N initially < cutoff | 27 of 284 | 8 of 61 | 5 of 103 | 6 of 30 | 8 of 90 |
| worsening (% , 95% CI) | (9.5%, 6.6% to 13.5%) | (13.1%, 6.8% to 23.8%) | (4.9%, 2.1% to 10.9%) | (20.0%, 9.5% to 37.3%) | (8.9%, 4.6% to 16.6%) |

MCID = minimal clinically important difference. <sup>a</sup>Cutoff threshold ≥ 10 for Patient Health Questionnaire-8.

### Multivariable Analysis of Factors Associated with Symptom Changes

As shown in Table 4, in adjusted analyses, compared to France, continuous scores for participants from the United States and the United Kingdom were 3.8 points (95% CI 1.7 to 5.9) and 2.9 points higher (95% CI 0.0 to 5.7); scores for Canada were higher but not statistically significant. Change scores were also significantly associated with pre-COVID PROMIS Physical Function 4a v1.0 scores and pre-COVID mental health service use; participants with pre-COVID-19 mental health service use had adjusted scores 3.7 points (95% CI 5.7 to 1.7) lower than other participants. Results were similar for odds of increasing by ≥ 1 MCID. Odds were approximately twice as high for participants from the United Kingdom (odds ratio 2.0, 95% CI 1.0 to 4.2) and United States (odds ratio 1.9, 95% CI 1.1 to 3.2) compared to France. Longer time since SSc diagnosis and overweight were also statistically significant. See Table 5.

**Table 4.** **Multivariable Analysis of Factors Associated with Change in Continuous Anxiety Symptom Scores Pre-COVID-19 to COVID-19**

| Variable | Unadjusted Regression | Adjusted Regression |
| --- | --- | --- |
|  | Coefficient<sup>a</sup> | Coefficient<sup>a</sup> |
|  | (95% Confidence Interval) | (95% Confidence Interval) |
| **Sociodemographic** |  |  |
| Age in years (continuous) | 0.02 (-0.05 to 0.08) | -0.05 (-0.12 to 0.02) |
| Male sex (reference = female) | -0.58 (-3.20 to 2.03) | -0.15 (-2.82 to 2.52) |
| Education in years (continuous) | 0.09 (-0.14 to 0.31) | 0.00 (-0.23 to 0.22) |
| Living alone (reference = living with others) | 1.24 (-0.92 to 3.40) | 1.45 (-0.66 to 3.56) |
| “Other” Race or ethnicity (reference = White) | -1.15 (-3.37 to 1.07) | 0.07 (-2.16 to 2.31) |
| Working part- or full-time (reference = not working) | 0.50 (-1.17 to 2.18) | -0.35 (-2.08 to 1.39) |
| Country (reference = France) |  |  |
| Canada | 1.22 (-0.98 to 3.43) | 1.64 (-0.59 to 3.89) |
| United Kingdom | 3.05 (0.27 to 5.84) | 2.88 (0.04 to 5.72) |
| United States | 3.81 (1.78 to 5.85) | 3.81 (1.69 to 5.94) |
| **Medical characteristics** |  |  |
| Body mass index (reference = underweight or normal) |  |  |
| Overweight | 0.96 (-1.03 to 2.95) | 1.27 (-0.71 to 3.24) |
| Obese | 0.81 (-1.52 to 3.15) | 2.15 (-0.23 to 4.54) |
| Time since diagnosis of SSc (continuous) | 0.11 (0.00 to 0.22) | 0.10 (-0.01 to 0.22) |
| Diffuse disease subtype (reference = limited or sine) | -0.65 (-2.36 to 1.06) | -0.69 (-2.50 to 1.11) |
| Presence of interstitial lung disease (reference = no) | 0.02 (-1.74 to 1.76) | 0.89 (-0.99 to 2.77) |
| Presence of any overlap syndrome (reference = no) | -0.13 (-2.13 to 1.87) | 0.18 (-1.84 to 2.20) |
| Immunosuppressant drug use (reference = no) | -0.34 (-2.01 to 1.33) | 0.35 (-1.51 to 2.21) |
| Pre-COVID-19 use of mental health services (reference = no) | -4.18 (-6.19 to -2.18) | -3.69 (-5.71 to -1.67) |
| Interference from breathing problems (continuous) | -0.54 (-0.83 to -0.26) | -0.28 (-0.63 to 0.07) |
| PROMIS Physical Function pre-COVID<br>(continuous) | 0.19 (0.10 to 0.29) | 0.14 (0.02 to 0.26) |
| **COVID-19 variables:** |  |  |
| Adequacy of financial resources = continuous | 0.23 (0.06 to 0.41) | 0.13 (-0.05 to 0.31) |

<sup>a</sup>Results based on imputed datasets. Based on assessment using via restricted cubic splines, there was no appreciable non-linearity.

**Table 5.** **Multivariable Analysis of Factors Associated with Change in of at least 1 MCID in Anxiety Symptom Scores Pre-COVID-19 to COVID-19**

| Variable | Unadjusted Odds Ratio<sup>a</sup><br>(95% Confidence Interval) | Adjusted Odds Ratio<sup>a</sup><br>(95% Confidence Interval) |
| --- | --- | --- |
| **Sociodemographic** |  |  |
| Age in years (continuous) | 1.01 (0.99 to 1.02) | 0.99 (0.97 to 1.01) |
| Male sex (reference = female) | 0.95 (0.53 to 1.72) | 0.99 (0.51 to 1.93) |
| Education in years (continuous) | 0.99 (0.94 to 1.04) | 0.98 (0.92 to 1.04) |
| Living alone (reference = living with others) | 1.13 (0.69 to 1.84) | 1.28 (0.74 to 2.22) |
| “Other” Race or ethnicity (reference = White) | 0.72 (0.44 to 1.20) | 0.88 (0.50 to 1.55) |
| Working part- or full-time (reference = not working) | 0.84 (0.57 to 1.22) | 0.73 (0.47 to 1.13) |
| Country (reference = France) |  |  |
| Canada | 1.18 (0.71 to 1.96) | 1.22 (0.69 to 2.13) |
| United Kingdom | 2.38 (1.23 to 4.59) | 2.02 (0.99 to 4.15) |
| United States | 1.96 (1.22 to 3.14) | 1.88 (1.10 to 3.19) |
| **Medical characteristics** |  |  |
| Body mass index (reference = underweight or normal) |  |  |
| Overweight | 1.59 (1.01 to 2.50) | 1.95 (1.18 to 3.24) |
| Obese | 1.13 (0.67 to 1.91) | 1.56 (0.86 to 2.86) |
| Time since diagnosis of SSc (continuous) | 1.05 (1.02 to 1.08) | 1.05 (1.02 to 1.09) |
| Diffuse disease subtype (reference = limited or sine) | 0.85 (0.58 to 1.25) | 0.83 (0.53 to 1.31) |
| Presence of interstitial lung disease (reference = no) | 1.06 (0.71 to 1.58) | 1.15 (0.71 to 1.86) |
| Presence of any overlap syndrome (reference = no) | 0.98 (0.62 to 1.53) | 0.95 (0.57 to 1.59) |
| Immunosuppressant drug use (reference = no) | 1.13 (0.78 to 1.65) | 1.43 (0.90 to 2.28) |
| Pre-COVID-19 use of mental health services (reference = no) | 0.51 (0.32 to 0.82) | 0.56 (0.34 to 0.94) |
| Interference from breathing problems (continuous) | 0.94 (0.88 to 1.00) | 0.97 (0.89 to 1.06) |
| PROMIS Physical Function pre-COVID<br>(continuous) | 1.02 (1.00 to 1.05) | 1.02 (0.99 to 1.06) |
| **COVID-19 variables:** |  |  |
| Adequacy of financial resources = continuous | 1.05 (1.01 to 1.10) | 1.05 (1.00 to 1.10) |

<sup>a</sup>Results based on imputed datasets.

Only country was statistically significantly associated with change in continuous depressive symptom scores; compared to participants from France, participants from Canada scored 1.5 points higher (95% CI 0.3 to 2.7), from the United Kingdom 1.8 points higher (95% CI 0.6 to 3.0), and from the United States 2.1 points higher (95% CI 0.5 to 3.7). See Supplementary Table 3. The odds ratio of an increase of ≥ 1 MCID was between 1.9 and 2.5 for the three countries, but only statistically significant for the United Kingdom (2.5, 95% CI 1.0 to 6.2). No other variables were statistically significant (Supplementary Table 4).

## DISCUSSION

We found that anxiety symptoms increased substantially compared to before the COVID-19 pandemic among vulnerable persons with a pre-existing medical condition, SSc, whereas depressive symptom changes were minimal. Overall, mean change on the PROMIS Anxiety 4a v1.0 was 4.9 points, greater than the MCID of 4 points. Approximately 50% of participants experienced an increase of ≥ 1 MCID. Results differed, however, by country. Anxiety symptoms increased by approximately 3 points among participants from France, 4 points among participants from Canada, 6 points among participants from the United Kingdom, and 7 points among participants from the United States. In multivariable analysis, compared to France, participants from the United Kingdom and United States scored approximately 3 and 4 points higher. Participants from the United Kingdom and the United States were also approximately twice as likely to have increased by ≥ 1 MCID, although this was not statistically significant for the United Kingdom. Another important finding was that people who used mental health services in the 3 months prior to their pre-COVID-19 assessment had adjusted anxiety symptom increases almost 4 points less than other participants.

Our study is one of the first to report mental health symptom changes during COVID-19 in a vulnerable population with a pre-existing medical condition and the first to compare symptom changes across countries. Compared to studies of university students, which suggest that depressive symptoms have increased by a small amount and anxiety minimally or not at all,^3,6^ we found that depressive symptoms changed minimally, but anxiety symptoms, on average, increased substantially. This may relate to the differential effect that COVID-19 is having on different segments of the population. University students may primarily be experiencing consequences of public health interventions, including interruption of academic programs, loss of work to support their studies, and reduced social connectedness. People with SSc and others with pre-existing medical conditions who are at risk of severe complications or death if infected likely perceive a greater threat from the virus than young adults of university age.

Increases in anxiety were larger in the United Kingdom and United States than in Canada and France. Comparing across countries is fraught with complexities. Nonetheless, the magnitude of the differences we found were surprisingly large. One possible explanation may relate to the coherence, or lack of coherence, of governmental and civil responses in the countries we studied. Indeed, editorials in the Lancet have described the American response as “inconsistent and incoherent”^25^ and the UK’s national response as “astonishingly haphazard.”^26^ France undertook some of the most restrictive measures internationally to attempt to reduce the spread of the virus,^27^ which may have reduced fear, relatively, among people vulnerable due to medical conditions. Canadian provinces were somewhat less restrictive but were generally consistent with a high level of political consensus on measures that have been taken.^28^

An important finding was that people who were using mental health services pre-COVID-19 had anxiety symptom change scores almost 1 MCID less than other participants. We do not know of evidence that people who have received mental health treatments are more resilient to adversity, meaning that they are able to adapt more successfully to negative or stressful situations. However, a recent systematic review and meta-analysis reported that psychological interventions designed to increase resilience do increase self-reported resilience.^29^ Many treatments for common mental illnesses include developing skills in structuring time, increasing the use of positive coping strategies, staying active even when not motivated, managing stress, and restructuring negatively distorted thought patterns, all of which would help to manage anxiety during a crisis like COVID-19.

There are limitations to consider. First, the SPIN Cohort is a convenience sample, although participant characteristics are similar to other large SSc cohorts.^8^ Second, participants complete questionnaires online, which may reduce generalizability. Third, it was not possible to capture and include local variables, such as the degree participants’ communities were affected or whether public health interventions were consistently followed in those communities. Nonetheless, data were collected at a time when social isolations were generally at their most conservative. Finally, different MCID values may be chosen. The 4-point MCID we used for anxiety symptoms was conservative; others have recommended MCIDs of 2 to 3 points,^30^ and it is possible that we may have underestimated the degree of patient-important change.

In sum, this was the first study to compare mental health symptoms prior to and during the COVID-19 outbreak. We found that anxiety symptoms increased substantially and that the magnitude was associated with country; increases were highest in the United States and United Kingdom and more moderate in France and Canada. There were minimal differences in depressive symptoms during COVID-19 compared to pre-COVID-19. An important finding was that people who received mental health care prior to COVID-19 experienced substantially smaller anxiety symptom increases than other participants. These findings, which differ from early reports of results from younger adults, underline that people at risk from medical complications in COVID-19 are also at risk of mental health problems and that there is a need for accessible mental health interventions that are designed to meet the specific needs of different populations.

## Contributors

BDT, LK, RSH, SP, SJB, JV, and ABenedetti were responsible for study concept and design. BDT, LK, RSH, SP, SH, ABourgeault, LT, SJB, JV, and ABenedetti were responsible for acquisition, analysis, or interpretation of data. LK, RSH, and ABenedetti were responsible for statistical analysis. BDT and RSH drafted the manuscript. All authors provided critical revision of the manuscript for important intellectual content, approved the final version, and agree to be accountable for all aspects of the work.

## Declaration of interests

All authors have completed the ICJME uniform disclosure form. Dr. Mouthon reported personal fees from Actelion/Johnson & Johnson, grants from LFB, non-financial support from Octapharma, and non-financial support from Grifols, all outside the submitted work. All other authors declare: no support from any organisation for the submitted work; no financial relationships with any organisations that might have an interest in the submitted work in the previous three years. All authors declare no other relationships or activities that could appear to have influenced the submitted work.

## Data Sharing

De-identified individual participant data with a data dictionary and analysis codes that were used to generate the results reported in this article will be made available upon request to the corresponding author and presentation of a methodologically sound proposal that is approved by the Scleroderma Patient-centered Intervention Network Data Access and Publications Committee. The study protocol and analysis plan are available at https://osf.io/kbncx/. Data will be available beginning 12 months after publication. Data requesters will need to sign a data transfer agreement.

## Acknowledgements

The study was supported by funding from the McGill Interdisciplinary Initiative in Infection and Immunity Emergency COVID-19 Research Fund; Scleroderma Canada, made possible by an educational grant for patient support programming from Boehringer Ingelheim; Scleroderma Society of Ontario; Scleroderma Manitoba; Scleroderma Atlantic; Scleroderma Australia; Scleroderma New South Wales; Scleroderma Victoria; Scleroderma Queensland; Scleroderma SASK; Scleroderma Association of BC; and Sclérodermie Québec. Drs. Thombs and Benedetti were supported by Fonds de recherche du Québec - Santé researcher salary awards, Dr. Henry was supported by a Mitacs postdoctoral fellowship award, and Mr. Harb was supported by a CIHR Canada Graduate Scholarship-Master’s award, all outside of the present work.

## Funding

McGill Interdisciplinary Initiative in Infection and Immunity Emergency COVID-19 Research Fund; Scleroderma Canada, made possible by an educational grant for patient support programming from Boehringer Ingelheim; Scleroderma Society of Ontario; Scleroderma Manitoba; Scleroderma Atlantic; Scleroderma Australia; Scleroderma New South Wales; Scleroderma Victoria; Scleroderma Queensland; Scleroderma SASK; Scleroderma Association of BC; and Sclérodermie Québec.

## Notes

### Clinical Protocols

https://osf.io/kbncx/

### Author Declarations

The SPIN (#MP-05-2013-150) and SPIN COVID-19 (#2021-2286) Cohorts were approved by the Research Ethics Committee of the Centre integre universitaire de sante et de services sociaux du Centre-Ouest-de-l'Ile-de-Montreal. The SPIN Cohort was also approved by ethics committees of SPIN sites.

